# SleepDepNet: A Multi-Task Transformer Framework for Assessing Sleep Quality and Depression Risk from Social Media Narratives

**DOI:** 10.1101/2025.04.17.25326012

**Authors:** Akshi Kumar, Saurabh Raj Sangwan, Aditi Sharma

## Abstract

The bidirectional relationship between sleep disturbances and depression presents a serious challenge for digital mental health research and intervention. This study introduces *SleepDepNet*, a transformer-based multi-task learning model designed to assess sleep quality and depressive sentiment simultaneously from user-generated narratives on Reddit. Leveraging a large, custom-labelled dataset drawn from subreddits such as *r/depression, r/sleep, r/mentalhealth*, and *r/insomnia*, SleepDepNet integrates attention mechanisms, sentiment and emotion analysis, and topic modelling to capture linguistic markers of emotional exhaustion and disordered sleep. The model achieves strong performance (F1-scores of 0.89 for sleep quality and 0.86 for depressive sentiment), while its attention-based interpretability supports transparent clinical insight. The proposed *SleepDepScore*, a unified metric derived from both tasks, offers a scalable approach to digital risk stratification and mental health triage. These results demonstrate SleepDepNet’s potential for real-world deployment in AI-driven mental health monitoring and personalized digital care.

## 1. Introduction

Sleep is an essential pillar of both physical and mental health, influencing a wide array of human functions such as cognitive performance, emotional regulation, and overall quality of life [1]. Sufficient and restorative sleep is critical for maintaining mental clarity, fostering resilience against stress, and ensuring physiological recovery. However, disturbances in sleep patterns—such as insomnia, irregular sleep cycles, or hypersomnia—often go together with mental health issues, especially depression [2]. For example, an individual experiencing chronic insomnia may begin to feel a loss of motivation, persistent sadness, or heightened anxiety, which are hallmarks of depressive episodes. Conversely, depression itself can disrupt sleep, creating a bidirectional relationship that compounds over time [3]. This complex interplay makes sleep and depression not just co-occurring issues but deeply intertwined phenomena requiring further exploration.

The significance of this relationship is underscored by statistics from the World Health Organization, which identifies depression as a leading cause of disability worldwide, affecting over 280 million people [4]. Among these, a significant proportion report sleep-related complaints, including difficulty falling asleep, frequent awakenings, or waking up unrefreshed. Clinical research has extensively documented this association, providing evidence of shared biological mechanisms, such as dysregulated circadian rhythms and altered neurotransmitter activity [5, 6]. However, while controlled clinical studies have yielded valuable insights, they often fail to capture the diversity and lived experiences of individuals in real-world settings.

In recent years, social media platforms like Reddit have emerged as unique venues for understanding health-related behaviours and concerns [7, 8]. Subreddits such as *r/depression, r/sleep, r/mentalhealth* and *r/insomnia* offer users a space to share candid narratives about their struggles with sleep, mental health, and daily life challenges. For instance, a user might post: *“I’ve been feeling exhausted lately. No matter how early I go to bed, I wake up feeling drained, and it’s making it harder to get through the day. I don’t know if it’s just stress or something deeper*.*”* Such posts provide rich, unfiltered accounts of personal experiences, often reflecting the subtle nuances of emotional and physical health that may not emerge in structured surveys or clinical interviews. These narratives offer a wealth of data that, if analyzed effectively, can deepen our understanding of how sleep and depression are interlinked. Despite the wealth of user-generated content available on platforms like Reddit, there is a notable gap in leveraging this data to study the intricate relationship between sleep and depression. Most existing research in this domain falls into one of two categories: (i) general sentiment analysis that fails to capture specific health-related dimensions, or (ii) studies relying on clinical datasets that, while precise, often lack the diversity and spontaneity of real-world narratives.

User-generated content on social media, by contrast, offers the opportunity to study authentic, varied expressions of sleep-related and depressive experiences at scale. However, analyzing such content poses its own set of challenges:

- *Noise and Variability:* Social media posts are informal, unstructured, and may contain irrelevant information.
- *Complexity of Language:* Expressions of sleep disturbances and depression can vary widely across individuals, with some using explicit terms (e.g., “insomnia”) and others relying on implicit cues (e.g., “feeling tired all the time”).
- *Multifaceted Interplay:* Posts often touch on multiple dimensions of well-being, requiring models capable of capturing these nuances.

To address these challenges, this study introduces SleepDepNet, a transformer-based, multi-task learning model designed to analyze user-generated narratives on Reddit. SleepDepNet performs two tasks simultaneously: (i) classifying sleep quality and (ii) detecting depressive sentiment. The model integrates advanced NLP techniques, including attention mechanisms, feature augmentation, and domain-specific embeddings, to address the limitations of traditional methods. Additionally, we introduce the SleepDepScore, a unified metric combining outputs from both tasks, enabling streamlined risk assessment and prioritization in real-world mental health applications. The problem statement is mathematical defined as follows:

Let D = {*d*_1_,*d*_2_, …,*d*_*n*_} represent a dataset of Reddit posts, where each post *d*_*i*_ consists of: **Text Content:** *T*_*i*_ = {*w*_1_, *w*_2_,…, *w*_*m*_} where *w*_*j*_ represents the *j-th* word in the post, **Metadata:** *M*_*i*_, including features such as timestamps, upvotes, and subreddit information.

Our objective is to design a multi-task learning model that simultaneously predicts:

i. **Sleep Quality Classification:** A binary label *y*_*i*_^(*s*)^ ∈ {0, 1}, where 0 indicates poor sleep quality and 1 indicates good sleep quality.
ii. **Depression Sentiment Analysis:** A sentiment label *y*_*i*_^(*d*)^ ∈ {―1, 0, 1},, where −1, 0, and 1 denote negative (depressive), neutral, and positive sentiment, respectively.

Additionally, we define the **SleepDepScore**, a unified index for combined risk assessment, as:

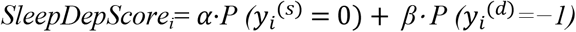

where:

- P (*y*_*i*_^(*s*)^) =0) is the probability of poor sleep quality.
- P (*y*_*i*_^(*d*)^= −1) is the probability of depressive sentiment.
- α and β are weights reflecting the relative importance of the two tasks in specific applications.

The multi-task learning objective is to minimize the weighted loss:

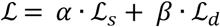

where ℒ_*s*_ and ℒ_*d*_are the individual task losses (e.g., cross-entropy loss). *α* and *β* are weights that balance the contributions of the two tasks.

This work makes the following key contributions:

- We propose SleepDepNet, a transformer-based multi-task learning model that simultaneously analyzes sleep quality and depressive sentiment in Reddit posts.
- We curate a custom dataset from Reddit subreddits like *r/depression, r/sleep, r/mentalhealth* and *r/insomnia*. The dataset is annotated with sleep quality labels and depressive sentiment categories, providing a valuable resource for future research.
- SleepDepNet study integrates sentiment analysis, emotion mapping, topic modelling, and linguistic features into BERT’s contextual embeddings, enhancing the model’s ability to capture domain-specific insights.
- SleepDepNet achieves superior performance compared to traditional baselines and transformer variants, with an F1-score of 0.89 for sleep quality prediction and 0.86 for depressive sentiment analysis.
- Through attention heatmaps, we provide insights into the model’s decision-making process, highlighting key linguistic features that correlate with sleep disturbances and depressive symptoms.
- Additionally, the introduction of SleepDepScore extends the model’s utility, offering a single, interpretable measure to prioritize interventions and assess combined sleep and emotional health risks.

By combining state-of-the-art NLP methods with real-world user data, SleepDepNet represents a significant step forward in understanding and addressing the complex interplay between sleep and mental health. The findings of this study have implications for both AI research and practical applications in healthcare, paving the way for more effective and accessible mental health interventions.

This paper is organized as follows: Section 1 introduces the background and significance of analyzing the interplay between sleep and mental health, motivating the need for advanced models like SleepDepNet. Section 2 reviews related literature, focusing on existing approaches to sleep quality prediction and depressive sentiment analysis, and highlights their limitations in handling unstructured data. Section 3 outlines the dataset creation process, detailing data collection, annotation, preprocessing, and distribution analysis, which serve as the foundation for model training. Section 4 describes the proposed SleepDepNet architecture, including feature extraction, multi-task learning framework, and optimization techniques. Section 5 presents experimental results, comparing SleepDepNet with baseline models and existing works, alongside detailed ablation studies and noise robustness evaluations. It also discusses model interpretability through attention visualizations and its implications for mental health applications. Section 6 discusses how SleepDepNet correlates sleep and depression through shared linguistic features, dataset analysis, and multi-task learning, validated by performance metrics, correlation analysis, and attention mechanisms. Finally, Section 7s concludes with key findings, limitations, and future research directions to enhance SleepDepNet’s scalability and applicability.

## 2. Literature Review

The detection of depression through text analysis has gained considerable attention, especially considering the increasing availability of user-generated content on platforms like Reddit [9]. Many studies have relied on pre-defined sentiment analysis methods to identify depressive language. For instance, Sharma and Sirts [10] explore depression-related posts on Reddit using rule-based approaches such as VADER, as well as transformer models like RoBERTa, to classify sentiment. While these models offer insights into depressive expressions, their scope was limited to sentiment polarity without searching into broader emotional or contextual modelling. In contrast, Tadesse et al. [11] applied machine learning models like bi grams with SVM, emphasizing the need for robust feature extraction for identifying depressive cues. However, these traditional models lack the contextual understanding required for complex user narratives. Kerasiotis et al. [12] advanced the field by combining transformer-based models such as DistilBERT with metadata and linguistic markers, achieving substantial gains in contextual representation and classification accuracy. Their work highlighted the importance of leveraging multiple transformer layers and weighted averages to enhance comprehension of textual patterns in depression-related posts. The integration of dropout layers and augmentation techniques further demonstrated the robustness of modern transformer-based architectures in handling class imbalance. Tejaswini et al. [13] extended the capabilities of text-based depression detection by integrating hybrid deep learning models such as FastText Convolutional Neural Networks with Long Short-Term Memory (FCL). Their approach combined the semantic representation power of FastText with the sequential learning ability of LSTM networks, demonstrating improved depression classification accuracy. Additionally, Uddin et al. [14] employed explainable artificial intelligence (XAI) techniques like LIME to enhance the interpretability of their LSTM-based models for depressive text detection, paving the way for more transparent mental health analytics.

In the domain of sleep quality prediction, wearable and mobile devices dominate the field [15, 16]. Lim et al. [17] highlighted the use of ecological momentary assessments (EMAs) combined with smart device data to understand lifestyle patterns influencing sleep quality. These studies focus on physiological data such as movement and heart rate, often ignoring linguistic data that could reveal user-reported sleep issues. Similarly, Zheng et al. [18] employed machine learning models such as artificial neural networks and random forests on structured data from the Pittsburgh Sleep Quality Index (PSQI). While these models achieved notable accuracy, they struggled to generalize beyond questionnaire-based inputs, limiting their application to broader populations.

Despite significant advancements in both depression detection and sleep quality prediction, few studies have explored their intersection. Most research isolates these domains, neglecting the shared underlying factors such as emotional stress and lifestyle patterns. Furthermore, datasets from platforms like Reddit often remain underutilized for integrated mental health analyses, despite their richness in linguistic and behavioural cues. Existing methods heavily favour structured datasets, such as EMAs or PSQI, over unstructured data from natural language. Additionally, many studies in depression detection prioritize identifying whether a post indicates depression or not, focusing on binary classification tasks such as “depression” versus “non-depression.” While this approach is effective in isolating posts indicative of depression, it often overlooks the finer gradations of emotional states. Sentiment polarity classification, which distinguishes between negative, neutral, and positive tones, provides valuable insights into the emotional nuances of depressive language and can enrich the understanding of user experiences beyond binary categorization. SleepDepNet bridges this gap by employing a transformer-based multi-task framework designed to simultaneously evaluate sleep quality and capture nuanced sentiment polarity, categorizing posts as negative, neutral, or positive. By integrating advanced NLP techniques such as attention mechanisms and emotion analysis, the model provides a detailed understanding of emotional states and their connection to sleep patterns. This approach surpasses the limitations of binary classification and rule-based models, offering a more granular and scalable solution for real-world mental health monitoring and intervention strategies.

## 3. Dataset Creation

A comprehensive and diverse dataset is essential for training robust models capable of understanding the complexities of sleep and mental health narratives. For this purpose, data was sourced from Reddit, a platform rich in user-generated content that reflects real-world experiences, challenges, and emotional states. This section outlines the steps taken to collect, annotate, and preprocess data, ensuring its suitability for training the SleepDepNet model. From sourcing posts across multiple subreddits to employing advanced preprocessing techniques, every stage was designed to capture the detailed linguistic and emotional patterns of discussions surrounding sleep and depression.

### 3.1. Data Collection

To create a comprehensive dataset for analyzing sleep and depression, we utilized the Python Reddit API Wrapper (PRAW), which provides efficient access to Reddit’s vast repository of user-generated content. Posts were collected from subreddits specifically focused on discussions about sleep patterns and mental health, including *r/depression, r/sleep, r/mentalhealth*, and *r/insomnia*. These subreddits were chosen because they are popular, highly active, and feature extensive discussions about sleep quality, mental health challenges, and coping strategies, making them rich sources of data for the proposed tasks. Each collected entry included multiple components:

- *Post Title:* A concise summary provided by the user, often highlighting their main concern or question. For instance, a title like *“Struggling with insomnia again”* succinctly frames the discussion around sleep difficulties.
- *Post Body:* The detailed narrative where users describe their experiences, emotions, and challenges. This is the primary text used for model training and provides valuable context for understanding user sentiment and sleep-related issues.
- *Comments:* Responses from other users, offering additional perspectives or support. While comments were not used for training in this study, they were collected for potential exploratory analyses or future expansions of the dataset.
- *Metadata:* Supplementary information about each post, including timestamps, subreddit type, and engagement metrics (e.g., number of upvotes, replies). These features provide additional context and can be useful for secondary analyses, such as tracking trends over time or assessing the community’s engagement with particular topics.

Over 50,000 posts were extracted, balanced across discussions related to sleep and depression. The balance was ensured by sampling posts evenly across the subreddits and filtering based on content relevance to the targeted themes. For example, posts focusing on unrelated topics, such as general life advice or technical questions, were excluded during the data collection process. This dataset forms the foundation for the SleepDepNet model and represents one of the largest annotated collections of Reddit posts focused on the interplay between sleep and depression. Its diversity in topics, language styles, and emotional expressions makes it an invaluable resource for understanding these complex phenomena through NLP.

### 3.2. Data Annotation

The dataset was annotated to support two key tasks: sleep quality classification and depressive sentiment analysis. These tasks were chosen to address the intertwined nature of sleep and depression, enabling a deeper understanding of the linguistic markers that characterize these phenomena.

For sleep quality classification, posts were categorized into two classes:

- **Good Sleep (1):** Posts indicating restful or satisfactory sleep experiences. For instance, a post like *“Finally managed to get 8 hours of sleep!”* reflects a positive sleep experience.
- **Poor Sleep (0):** Posts highlighting sleep difficulties, such as *“I can’t sleep no matter what I do*.*”* These posts often describe struggles like insomnia, frequent awakenings, or inadequate rest.

For depressive sentiment analysis, categorizing posts into three sentiment classes—negative, neutral, and positive—provides a nuanced understanding of the emotional spectrum in user narratives:

- **Negative Sentiment (−1):** Posts that convey depressive symptoms or negative emotions, such as *“I feel hopeless and exhausted*.*”* These posts often include expressions of sadness, fatigue, or despair. These posts are essential for identifying individuals experiencing acute depressive states, enabling targeted interventions.
- **Neutral Sentiment (0):** Posts that neither strongly reflect depressive symptoms nor positive emotions, e.g., *“Tried a new sleep schedule, not sure if it works yet*.*”* These posts tend to be descriptive or factual without an overt emotional tone. This category captures posts that do not align with depressive or optimistic sentiments, providing balance and reducing ambiguity in the dataset.
- **Positive Sentiment (1):** Posts showing improvement or a positive outlook, for example, *“Woke up feeling great today after a long time*.*”* Including this category is particularly valuable for understanding progress, resilience, and coping mechanisms among users. By identifying positive sentiment, the analysis highlights opportunities to reinforce recovery strategies and support mental well-being.

This three-tier classification ensures that the model captures the complexity of depressive sentiment, moving beyond binary sentiment analysis to provide a more comprehensive understanding of user expressions. It also facilitates actionable insights for mental health professionals to tailor interventions based on the emotional state of individuals.

The annotation process was conducted in two stages to ensure accuracy and scalability. Initially, posts were labelled using a combination of keyword-based rules and sentiment lexicons. For instance, keywords like *“tired,” “awake all night,”* and *“insomnia”* were used to classify poor sleep, while terms like *“refreshed”* and *“well-rested”* suggested good sleep. Similarly, sentiment lexicons helped categorize posts into sentiment classes based on emotional tone. To refine the initial annotations, ChatGPT was employed to review and adjust labels for 10,000 randomly selected posts. ChatGPT was provided with context about the classification tasks and guidelines for interpreting the text. For example, it was instructed to distinguish between posts explicitly mentioning depressive symptoms and those with neutral or unrelated content. ChatGPT’s outputs were further cross verified for consistency and adherence to the annotation criteria. This hybrid approach of rule-based classification followed by ChatGPT-assisted validation ensured that the dataset was both large-scale and highly accurate. The annotated dataset is now well-suited for training the SleepDepNet model, offering a reliable foundation for analyzing the intricate connections between sleep quality and depressive symptoms.

### 3.3. Analysis of Dataset Distribution

The dataset comprises a total of 50,000 Reddit posts, meticulously categorized into sleep quality and depressive sentiment classes. These statistics provide a comprehensive view of the dataset’s structure and highlight the linguistic and emotional trends prevalent in discussions related to sleep and mental health. (Table 1)

**Table 1.** Dataset Distribution.

| Category | Number of Posts |
| --- | --- |
| Good Sleep | 15,000 |
| Poor Sleep | 35,000 |
| Negative Sentiment | 25,000 |
| Neutral Sentiment | 15,000 |
| Positive Sentiment | 10,000 |

#### 3.3.1. Sleep Quality Classification

- **Good Sleep (15**,**000 posts)**: Comprising 30% of the dataset, this category includes posts where users report restful and satisfying sleep experiences. For example, posts like *“Finally managed to sleep uninterrupted for 8 hours last night!”* reflect positive sleep outcomes. While this category is smaller compared to poor sleep, it offers valuable insights into the language used by individuals with healthy sleep habits.
- **Poor Sleep (35**,**000 posts)**: Representing the remaining 70% of posts in this category, these narratives highlight significant sleep difficulties such as insomnia, interrupted sleep, or chronic fatigue. Posts like *“I haven’t been able to sleep for days, and it’s taking a toll on my mood and energy levels*.*”* dominate this category, emphasizing the widespread prevalence of sleep issues in the sampled data.

#### 3.3.2. Depressive Sentiment Analysis

- **Negative Sentiment (25**,**000 posts)**: Accounting for 50% of the dataset, these posts express depressive symptoms, including sadness, hopelessness, and emotional exhaustion. Examples include *“I feel so empty and drained, like nothing matters anymore*.*”* This category underscores the significant emotional burden associated with poor mental health.
- **Neutral Sentiment (15**,**000 posts)**: Encompassing 30% of the dataset, neutral posts are largely factual or exploratory, such as *“I’ve been experimenting with new sleep routines; not sure if they’re working yet*.*”* These posts provide a balanced context and serve as a baseline for sentiment analysis.
- **Positive Sentiment (10**,**000 posts)**: The smallest subset (20%) includes posts reflecting optimism or recovery, such as *“Finally had a good night’s sleep and feel much better today*.*”* These posts offer critical insights into patterns of improvement and coping mechanisms.

The dataset reveals several key observations that highlight its utility and alignment with real-world trends in sleep and mental health discussions. Firstly, there is an imbalance in sleep quality labels, with significantly more posts classified as *“Poor Sleep”* (70%) compared to *“Good Sleep”* (30%). This disparity mirrors real-world patterns, where sleep disturbances like insomnia and irregular sleep cycles are more commonly reported than experiences of restful and restorative sleep. Such a skewed distribution is expected, given the focus of the sampled subreddits, which attract users seeking support or solutions for sleep-related challenges. Secondly, the prevalence of negative sentiment in the dataset is notable, accounting for 50% of the depressive sentiment analysis labels. This reflects the emotional tone commonly found in subreddits like *r/depression*, where users frequently share struggles with mental health and emotional well-being. The large proportion of “Negative Sentiment” posts underscores the importance of addressing mental health challenges in online communities and emphasizes the relevance of developing models like SleepDepNet to analyze and respond to such content effectively. Despite the dominance of negative sentiment, the dataset also includes a meaningful representation of neutral (30%) and positive (20%) sentiments, which ensures a more nuanced and comprehensive understanding of the emotional spectrum. Neutral posts, often factual or exploratory, provide a baseline for analysis, while positive posts offer insights into recovery, improvement, and coping mechanisms. This diversity in sentiment labels enriches the dataset, enabling SleepDepNet to differentiate between varied emotional tones and identify patterns of resilience and positivity alongside challenges. Collectively, these observations underscore the richness and diversity of the dataset, making it a robust resource for training and evaluating SleepDepNet. The balance and distribution across sleep quality and sentiment categories ensure that the model is exposed to a wide array of linguistic and emotional patterns. This exposure enhances the model’s ability to make accurate predictions and generate actionable insights, thereby contributing to advancements in the domain of sleep and mental health.

### 3.4. Data Preprocessing

To prepare the dataset for training, a comprehensive preprocessing pipeline was implemented to clean, standardize, and transform the raw text data into a format suitable for the SleepDepNet model. The first step involved text cleaning, which removed URLs, mentions (e.g., @username), hashtags, and special characters, ensuring that irrelevant elements were eliminated from the posts. All text was converted to lowercase to maintain uniformity and prevent case sensitivity issues. Subsequently, the WordPiece tokenizer, integral to transformer models like BERT, was used to tokenize the text. This tokenizer breaks words into sub word units, effectively handling out-of-vocabulary words and improving representation for rare or compound terms. Following text cleaning, stop word removal was applied to reduce noise by eliminating commonly used words such as “the” and “and” that do not contribute significant meaning. However, domain-specific stop words relevant to sleep and depression, such as “tired,” “sad,” and “awake,” were retained to preserve critical contextual signals. To further standardize linguistic variations, lemmatization was performed, reducing words to their base forms. For instance, variations like “sleeping” and “sleeps” were converted to “sleep,” and “tiredness” to “tired,” ensuring semantic consistency while minimizing redundancy. Given the variability in post lengths, padding and truncation were applied to ensure all input sequences conformed to a fixed length of 128 tokens. Truncation was used to shorten excessively long posts, while padding added zeros to shorter posts to maintain uniform sequence lengths across the dataset. This step was crucial for enabling efficient batch processing during model training and inference. Finally, embedding preparation was carried out using the pre-trained BERT tokenizer, which transformed the cleaned and tokenized text into high-dimensional vector representations. These embeddings capture the semantic and syntactic nuances of the text, forming the input features for the SleepDepNet model. By implementing these preprocessing steps in a systematic and balanced manner, the dataset was transformed into a standardized and enriched format, ensuring that noise was minimized while meaningful linguistic and contextual features were retained.

## 4. Model Architecture

SleepDepNet is a robust multi-task transformer-based model designed to classify sleep quality and analyze depressive sentiment in Reddit posts. The architecture leverages a shared transformer encoder, feature extraction and augmentation techniques, and attention mechanisms for enhanced interpretability and performance. It integrates advanced techniques like emotion mapping, topic modelling, and correlation analysis to achieve precise and actionable insights.

### 4.1 Shared Transformer Encoder

At the core of SleepDepNet is a fine-tuned BERT (Bidirectional Encoder Representations from Transformers) model [19], which serves as the shared encoder. BERT is a pre-trained transformer architecture that captures contextual relationships between words in a sequence, making it well-suited for tasks involving complex natural language understanding. The model leverages its contextual richness to accurately capture the nuanced relationships between words and their surrounding context. For example, it differentiates between the phrase *“I’m tired of feeling this way”* (indicating depressive sentiment) and *“I’m tired after work”* (a neutral expression).

- **Input Representation**: The input to the model consists of the concatenated post title and post body from the Reddit dataset. Metadata such as timestamps, upvotes, and subreddit information is also included to provide contextual signals These text components are tokenized using BERT’s WordPiece tokenizer, which splits the text into sub word units. This ensures that even rare or misspelled words are adequately represented. The input sequence is represented as (equation 1):

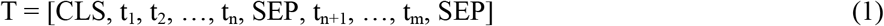

Here, CLS is a special token representing the entire input sequence, while SEP separates the title and body.
- **Shared Representation Vector**: The tokenized input is fed into BERT, which generates contextual embeddings for each token in the sequence. The embedding corresponding to the CLS token is extracted as the shared representation vector (equation 2):

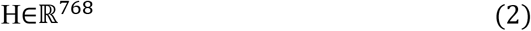

This vector encapsulates the semantic and contextual information of the entire input text.

### 4.2. Feature Extraction and Augmentation

To enhance BERT’s contextual embeddings, a Feature Augmentation Layer processes Reddit posts using complementary techniques, integrating linguistic, semantic, and contextual signals. These additional features enable the SleepDepNet model to capture a richer representation of the data, improving predictive accuracy and interpretability for sleep quality classification and depressive sentiment analysis.

#### 4.2.1. Sentiment and Emotion Analysis

To enhance the SleepDepNet model’s capacity to capture and interpret emotional nuances in Reddit posts, a detailed sentiment and emotion analysis pipeline was integrated. This step aimed to preprocess and classify emotional content, thereby enriching the feature representations utilized by the model for downstream tasks.

##### a. Sentiment Analysis with VADER

The Valence Aware Dictionary and Sentiment Reasoner (VADER) [20] was applied as an initial tool to determine the sentiment polarity of posts. VADER assigns sentiment scores in three categories: positive, neutral, and negative. For a given post *P*, the VADER model evaluates the valence of each word *w*_*i*_and computes an overall sentiment score *S(P)* as a weighted sum of individual word scores (equation 3):

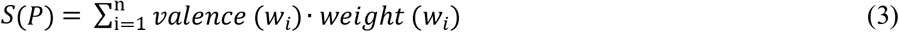

where *valence* (*w*_*i*_) is the sentiment intensity of word *w*_*i*_ and *weight* (*w*_*i*_) adjusts for emphasis (e.g., capitalization, punctuation). Posts with *S*(*P*) > τ_1_ were classified as positive, those with *S*(*P*) < τ_2_ as negative, and values in between as neutral, where τ_1_ and τ_2_ are predefined thresholds.

##### b. Refined Sentiment Classification with RoBERTa

While VADER provides a lexicon-based baseline, it may struggle with nuanced or contextual sentiment. To address this, a fine-tuned RoBERTa (Robustly Optimized BERT Approach) model [21] was employed. RoBERTa uses transformer layers to generate contextual embeddings for posts and predicts sentiment with higher precision for complex and figurative language. Given a tokenized input sequence T = [CLS, t_1_, t_2_, …, t_n_, SEP], RoBERTa processes the sequence and produces a contextual embedding *H*_CLS_ for the classification token (equation 4):

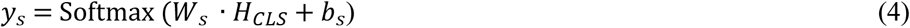

where *W*_*s*_ ∈ ℝ^3×*d*^ is the weight matrix, *b*_*s*_∈ ℝ^3^ is the bias vector, and *d* is the dimensionality of the hidden state. The output *y*^(*s*)^represents probabilities for the sentiment classes: positive, neutral, and negative. Fine-tuning RoBERTa on sentiment-specific datasets enabled the model to adapt to the unique language used in Reddit posts.

##### c. Emotion Mapping with NRC Emotion Lexicon

Beyond sentiment, the NRC Emotion Lexicon [22] was employed to categorize posts into specific emotional states such as sadness, frustration, or anger. Each word in a post *P* was matched to predefined emotion categories *ε* = {sadness, joy, anger, …} using the lexicon (equation 5):

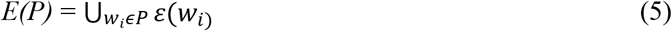

where *ε*(*w*_*i*)_returns the set of emotions associated with word *w*_*i*_. For example, the phrase *“I feel so drained and hopeless”* would map to *ε* = {sadness, frustration}. Posts with multiple emotional states were assigned a composite label, reflecting the complexity of user narratives.

##### d. Integration into Feature Representations

The outputs of these analyses informed the feature representations for the classification tasks. For instance:

- Posts with high negative sentiment scores from VADER or RoBERTa were correlated with depressive language features.
- Emotional categories derived from the NRC Emotion Lexicon added granularity to the contextual embeddings by emphasizing relevant emotional dimensions.

For example, the post *“I can’t sleep at night and feel completely hopeless”* might produce the following enriched representation:

- **Sentiment**: Negative (VADER score = −0.85, RoBERTa probability = 0.92 for negative class).
- **Emotion**: Sadness, frustration (based on lexicon mappings).

These analyses provided depth to the model’s contextual understanding, ensuring that SleepDepNet could effectively leverage emotional and sentiment-based insights to improve classification performance. By combining lexicon-based and transformer-based methods, the sentiment and emotion analysis pipeline bridged the gap between simple rule-based systems and advanced contextual embeddings.

#### 4.2.2. Topic Modelling and Correlation Analysis

To delve deeper into the thematic patterns and relationships within Reddit discussions, topic modelling and correlation analysis were conducted. These techniques helped identify recurring themes and explore the interplay between sleep-related issues and depressive language patterns.

##### a. Topic Modelling with Latent Dirichlet Allocation (LDA)

LDA [23], a generative probabilistic model, was used to uncover latent themes in the textual data. LDA assumes that each post is a mixture of topics, and each topic is a distribution over words. Given a set of *N* posts, LDA represents the probability of a word *w* belonging to a topic *z* within a post *d* as (equation 6):

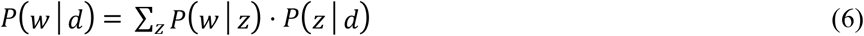

where:

- *P* (*w*|*z*) is the probability of word *w* given topic *z*,
- *P* (*z*|*d*) is the probability of topic *z* given post *d*.

The LDA model identified two primary themes in the dataset: causes of poor sleep and coping mechanisms for depression. To visualize these topics, word clouds were generated from the most frequently occurring terms associated with each theme (Fig. 1 and 2).

**Fig. 1.**
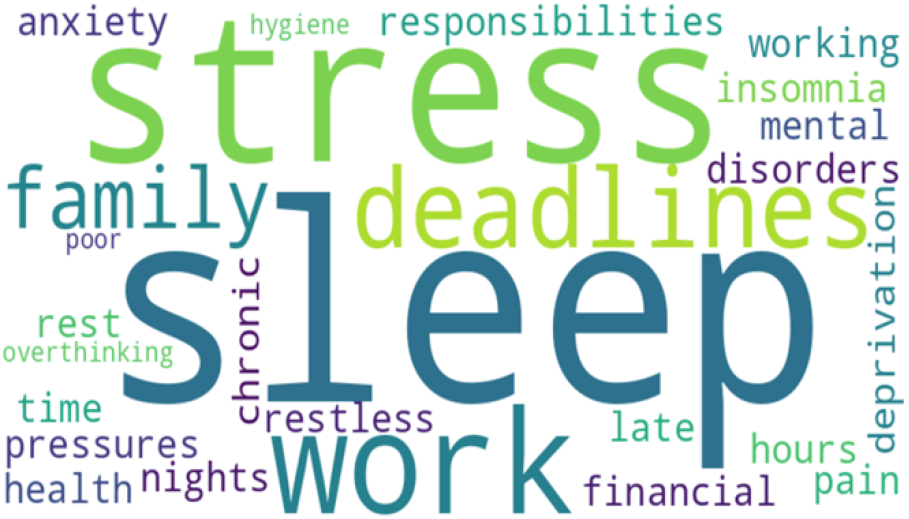
Word Cloud for Causes of Poor Sleep

**Fig. 2.**
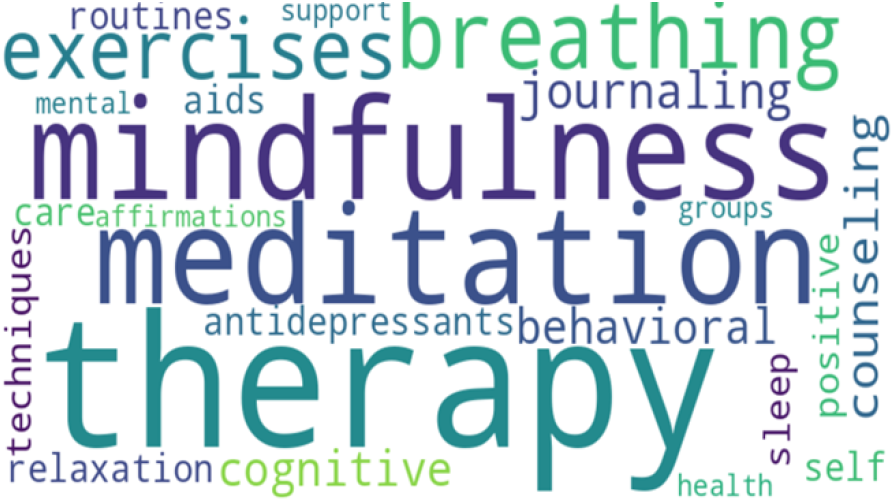
Word Cloud for Coping Mechanisms for Depression

These word clouds highlight critical terms and phrases that provide insight into the discussions within the dataset.

##### b. Correlation Analysis

Building upon the insights from topic modelling, a correlation analysis was conducted to quantify relationships between specific terms related to sleep and depression. Co-occurrence frequencies were calculated for term pairs (e.g., *“insomnia”* and *“hopeless”*) across posts, followed by the application of Pearson correlation [24] to measure the strength of associations. The Pearson correlation coefficient *r* was computed using (equation 7):

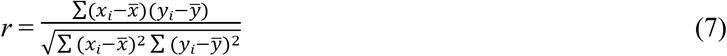

where *x*_*i*_ and *y*_*i*_ are the frequencies of terms in individual posts, and *x* and *y* are their respective mean frequencies. The findings were as follows:

- **Strong Correlations**: Terms such as *“insomnia”* and *“hopeless”* exhibited high correlation coefficients (*r* > 0.8), suggesting a frequent co-occurrence in user posts discussing emotional struggles and sleep difficulties.
- **Moderate Correlations**: Terms like *“sleepless”* and *“exhausted”* showed moderate correlations (0.6 ≤ *r* < 0.8), emphasizing their relevance to fatigue and emotional distress.
- **Weaker Correlations**: Some terms, like *“restless”* and *“depressed”*, exhibited lower correlations (*r* < 0.6) but still indicated a meaningful relationship.

To better understand these relationships, a heatmap was generated (Fig.3) to display the Pearson correlation coefficients between term pairs.

The heatmap highlights the correlation strengths, with darker shades indicating stronger relationships. For example:

**Fig. 3.**
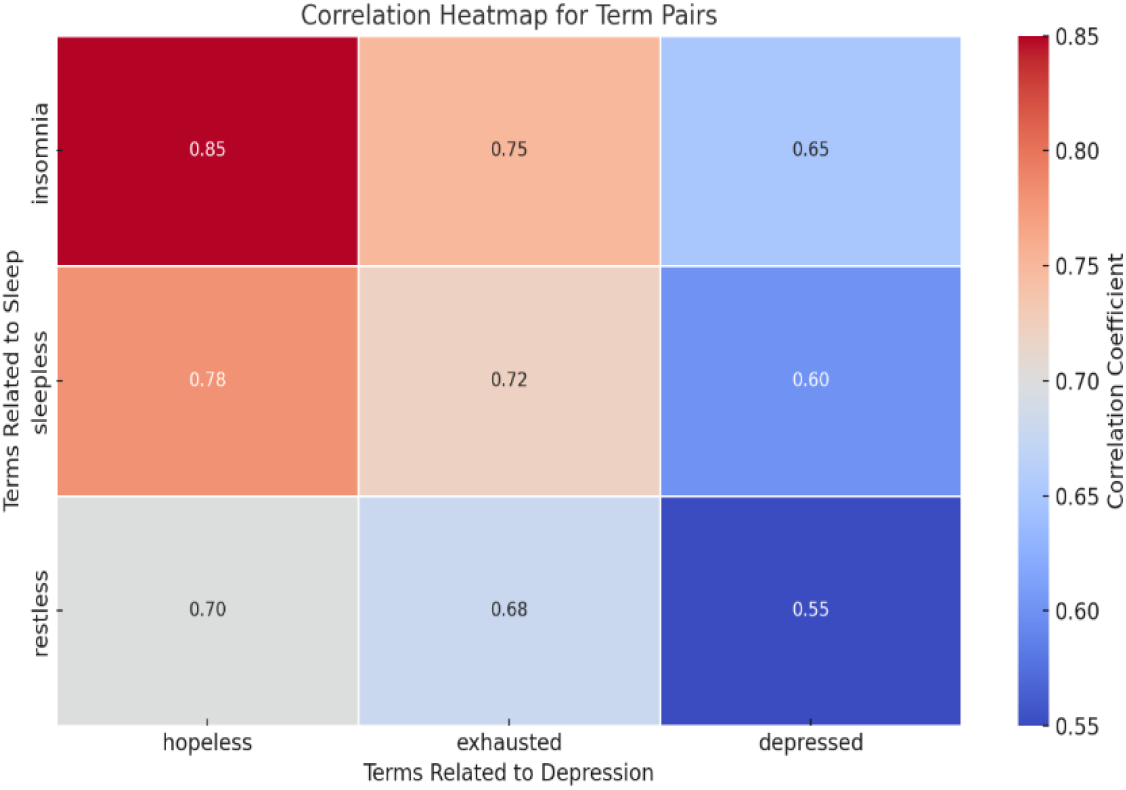
Correlation Heatmap for Term Pairs

- High Correlation: *“Insomnia”* and *“hopeless”* (r=0.85).
- Moderate Correlation: *“Sleepless”* and *“exhausted”* (r=0.72).
- Low Correlation: *“Restless”* and *“depressed”* (r=0.55).

The correlation analysis confirms the interplay between sleep difficulties and depressive expressions, validating the importance of analyzing these terms together in understanding user narratives. These insights were used to enhance the feature representations in SleepDepNet. The integration of LDA topic modelling and correlation analysis significantly enriched the SleepDepNet pipeline, adding layers of depth and contextual understanding. Through topic modelling, the pipeline gained thematic insights by uncovering macro-level patterns of recurring issues and solutions within the data. This informed the model about key domains of user concern, such as the causes of poor sleep and coping mechanisms for depression, ensuring that the model’s focus aligned with user-relevant topics. Meanwhile, correlation analysis enhanced the model’s ability to capture nuanced relationships between specific terms, such as the frequent co-occurrence of *“insomnia”* with *“hopeless”* or *“restless”* with *“exhausted*.*”* These contextual relationships allowed SleepDepNet to associate linguistic patterns with underlying emotional states more effectively, ultimately improving its predictive accuracy and interpretability. By combining these approaches, the pipeline bridged high-level thematic understanding with detailed linguistic correlations, resulting in a robust framework for analyzing sleep and depression narratives. For example, a post stating *“I can’t fall asleep because I’m constantly worried about my work deadlines”* would contribute to the stress-related causes of poor sleep topic and highlight a correlation between “can’t sleep” and “worried.” This enriched contextual understanding allowed SleepDepNet to make more accurate predictions and offered actionable insights into the connections between sleep and depression.

#### 4.2.3. Additional Linguistic Features

TF-IDF (Term Frequency-Inverse Document Frequency) [25] was used to identify key terms in the discussions related to sleep and depression. By evaluating the importance of words relative to the corpus, this technique highlighted terms that were frequently discussed within specific contexts, such as *“insomnia”* or *“hopeless,”* while down-weighting commonly occurring but less informative words like *“and”* or *“the*.*”* This approach ensured that the extracted features were representative of the unique linguistic patterns within the dataset. Word2Vec [26], a neural embedding technique, was employed to capture semantic relationships between words. By representing words as dense vectors in a continuous vector space, Word2Vec enabled the model to understand latent patterns and associations in user language. For example, words like *“fatigue”* and *“exhaustion”* might appear closer in the vector space, reflecting their contextual similarity in discussions about depression. In addition to these advanced representation techniques, specific features related to sleep and depressive language were extracted from the text. Terms like *“insomnia,” “awake all night,”* and *“eight hours of sleep”* were identified as indicators of sleep quality. Similarly, phrases such as *“hopeless,” “exhausted,”* and *“drained”* were highlighted to capture depressive symptoms and emotional states. By focusing on these domain-specific features, the model was primed to recognize patterns and themes relevant to sleep and mental health.

#### 4.2.4. Integration with BERT

The augmentation of contextual Embeddings leverages features derived from sentiment analysis, emotion mapping, topic modelling, and linguistic signals to enrich BERT’s contextual embeddings. This approach ensures that both semantic and syntactic information are integrated into the input representation, enhancing the data’s utility for downstream tasks. For instance, while BERT effectively captures the context in a phrase like *“I can’t sleep because I’m stressed,”* the augmented features add layers of understanding by highlighting sentiment (negative), emotions (frustration), and thematic relevance (stress as a cause of poor sleep). These additional signals strengthen the model’s comprehension of domain-specific nuances. The rationale for feature augmentation lies in addressing BERT’s potential limitations in identifying domain-specific insights, such as emotional categories or thematic patterns. Although BERT excels in capturing linguistic complexity, it may overlook critical task-specific details. Feature augmentation bridges this gap by incorporating domain-relevant and task-specific knowledge to create a comprehensive input representation. By integrating lexicon-based tools like VADER and NRC Emotion Lexicon, probabilistic methods like LDA, embedding-based techniques such as TF-IDF and Word2Vec and RoBERTa for refined sentiment analysis, the model gains a multi-dimensional perspective on user narratives. Thus, the outputs from sentiment analysis, emotion mapping, topic modelling, and linguistic features are combined into a unified vector representation. These features are concatenated with BERT’s contextual embeddings from the shared transformer encoder. This multi-faceted augmentation process ensures that the input features are robust, domain-aware, and tailored for analyzing complex relationships within the data. It provides a strong foundation for the downstream layers of SleepDepNet, enabling the model to perform highly accurate and interpretable predictions in tasks such as sleep quality classification and depressive sentiment analysis.

### 4.3. BiLSTM with Attention

To complement the transformer-based architecture, a Bidirectional Long Short-Term Memory (BiLSTM) network with an attention mechanism [27] was incorporated. BiLSTM is particularly effective in capturing sequential dependencies, making it ideal for processing the temporal and contextual flow of user narratives. Unlike standard LSTMs, BiLSTMs process text in both forward and backward directions, generating two hidden states for each input. These hidden states are concatenated to form a comprehensive representation of each word in its context (equation 8):

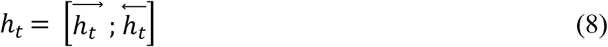

where 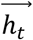 and 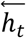 are the forward and backward hidden states, respectively.

To further enhance interpretability and focus on the most relevant text segments, an attention mechanism was added. The attention layer assigns a weight *α*_*t*_ to each hidden state, quantifying its importance in predicting the target labels. The final context vector *c* is computed as a weighted sum of the hidden states (equation 9):

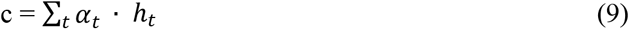

where 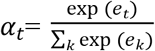, and *e*_*t*_ is a learned alignment score. This mechanism ensures that the model emphasizes critical words or phrases, such as *“can’t sleep”* or *“feeling drained,”* which are strongly indicative of sleep disturbances or depressive symptoms.

### 4.4. Task-Specific Layers

The task-specific layers of SleepDepNet utilize a shared representation enriched with sequential features from BiLSTM and contextual embeddings from BERT. Fully connected layers then project this representation into task-specific spaces, enabling accurate predictions for sleep quality classification and depressive sentiment analysis.

#### 4.4.1. Sleep Quality Classification Head

For the sleep quality classification task, a fully connected layer is applied to the shared representation. This layer projects the 768-dimensional vector into a 2-dimensional space, representing probabilities for the two classes: Good Sleep (1) and Poor Sleep (0). The output is calculated as (equation 10):

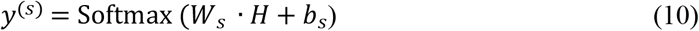

Where:

- *W*_*s*_ ∈ ℝ^2×768^ is the weight matrix for the sleep classification task.
- *b*_*s*_∈ ℝ^2^ is the bias vector.
- *y*^(*s*)^contains the probabilities for each class.

#### 4.4.2. Depressive Sentiment Analysis Head

For the depressive sentiment analysis task, a separate fully connected layer projects the shared representation into a 3-dimensional space, corresponding to the sentiment classes: Negative (−1), Neutral (0), and Positive (1). The output is computed as (equation 11):

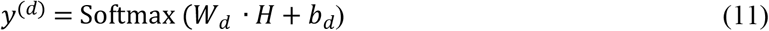

Where:

- *W*_*d*_ ∈ ℝ^3×768^ is the weight matrix for the sentiment analysis task.
- *b*_*d*_∈ℝ^3^ is the bias vector.
- *y*^(*d*)^contains the probabilities for each sentiment class.

Both task-specific heads leverage the shared representation H, allowing the model to optimize for both tasks simultaneously.

### 4.5. Interpretability via Attention Maps

To improve the interpretability of SleepDepNet’s predictions, an attention mechanism [28] was incorporated into the architecture. This mechanism generates attention maps that visualize the significance of specific words or phrases in the input text, highlighting the tokens that most influence the model’s decisions for each task. For instance, in the sleep quality classification task, the model may assign higher attention weights to terms like *“insomnia”* or *“awake all night”* when predicting poor sleep quality. Similarly, in depressive sentiment analysis, phrases such as *“feeling hopeless”* or *“I’m exhausted”* might receive greater focus when identifying negative sentiment. By offering a clear visualization of the decision-making process, the attention mechanism enhances transparency and provides actionable insights, making the model’s reasoning more interpretable for both researchers and practitioners.

### 4.6. Multi-Task Learning and Optimization

SleepDepNet employs a multi-task learning framework, optimizing both tasks simultaneously using a shared encoder. The overall loss function is a weighted combination of the task-specific losses (equation 12):

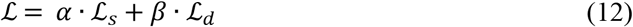

Where:

- ℒ_*s*_: Binary cross-entropy loss for sleep quality classification.
- ℒ_*d*_: Categorical cross-entropy loss for depressive sentiment analysis.
- *α* and *β*: Hyperparameters controlling the contributions of each task.

The optimization of SleepDepNet is performed using the AdamW optimizer, a variant of Adam that incorporates weight decay to reduce overfitting and improve generalization. To ensure a stable and effective learning process, a learning rate schedule is employed, beginning with a warm-up phase to gradually increase the learning rate at the start of training, followed by linear decay to decrease the learning rate as training progresses. This approach helps the model converge effectively while avoiding instability during the initial stages of training. Additionally, gradient clipping is applied to cap the magnitude of gradients, preventing exploding gradients that could disrupt the optimization process. To further improve generalization and reduce the risk of overfitting, the model incorporates several regularization techniques. Dropout is applied to the task-specific layers with a rate of 0.1, randomly dropping a fraction of connections during training to discourage the model from relying too heavily on specific features. Within the BERT architecture, Layer Normalization is utilized to stabilize training and ensure smooth convergence by normalizing the inputs to each layer, maintaining consistent gradients. Finally, an early stopping criterion is employed, halting training when validation performance stops improving, thereby avoiding overfitting to the training data. These optimization and regularization strategies collectively ensure that SleepDepNet achieves robust and reliable performance across its tasks.

### 4.7. Unified Score for Risk Assessment

To complement the task-specific outputs, SleepDepNet introduces the SleepDepScore, a composite metric designed to integrate the outputs of both tasks into a single risk index. The unified score is particularly beneficial for ranking user narratives or identifying high-risk cases for intervention. The score is computed as (equation 12):

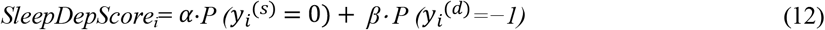

where:

- P (*y*_*i*_^(*s*)^) =0) is the probability of poor sleep quality.
- P (*y*_*i*_^(*d*)^= −1) is the probability of depressive sentiment.
- α and β are weights reflecting the relative importance of the two tasks in specific applications.

This approach not only enhances interpretability but also provides a practical metric for real-world applications, such as mental health monitoring systems

Below is a conceptual diagram illustrating the architecture of SleepDepNet (Fig.4):

**Fig. 4.**
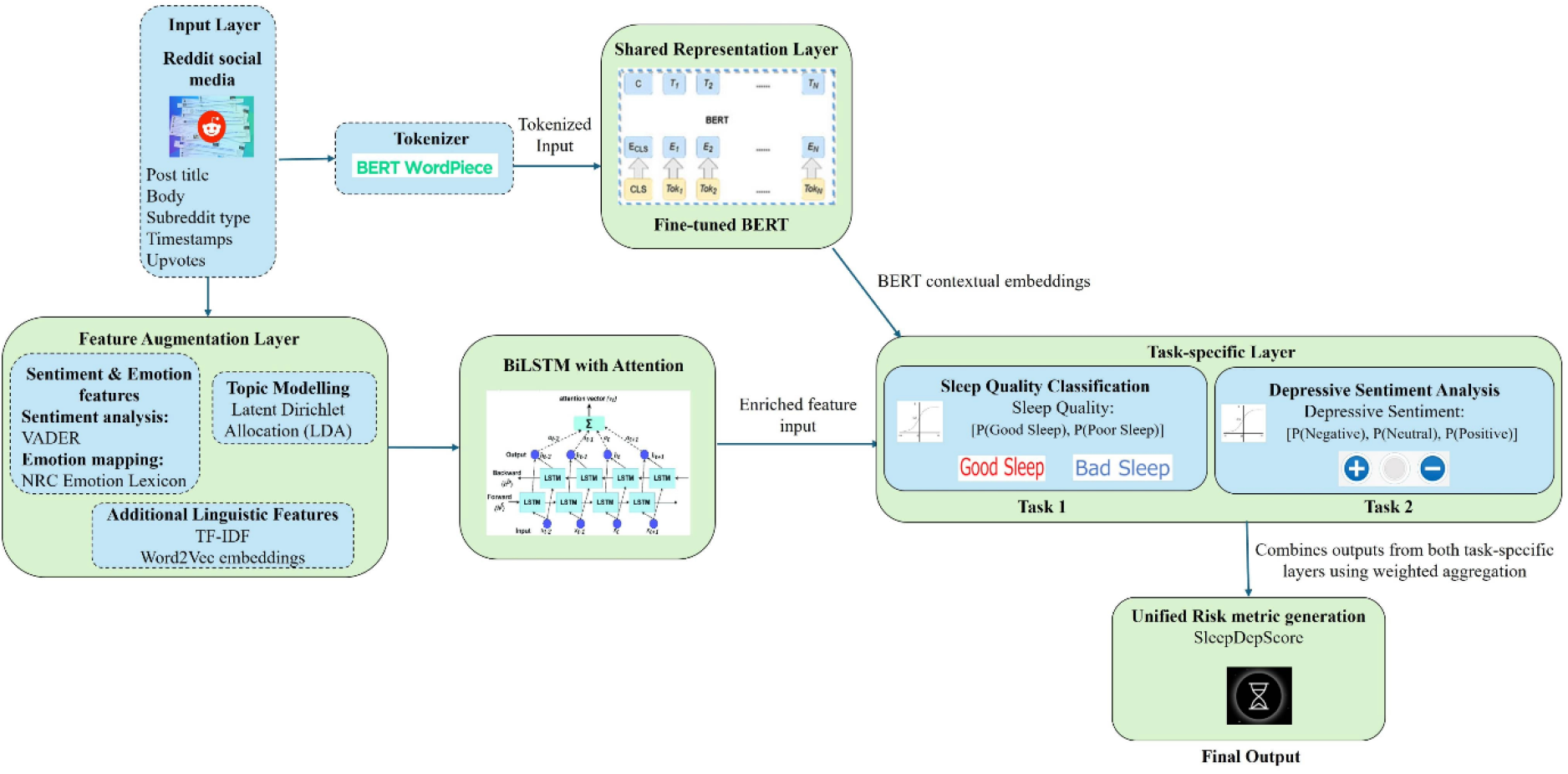
SleepDepNet Architecture

## 5. Implementation and Results

The Implementation and Results section provides an in-depth examination of SleepDepNet’s development, setup, evaluation, and performance across various metrics and configurations. It systematically evaluates the contributions of individual model components, compares performance against baseline and existing methods, and showcases the robustness of SleepDepNet through comprehensive experiments and visualizations.

### 5.1 Setup

The implementation of SleepDepNet was conducted using a robust computational setup designed for efficiency and scalability. The hardware environment included a NVIDIA Tesla V100 GPU with 32 GB of VRAM for accelerated training, an Intel Xeon Gold 6230R CPU, and 256 GB of RAM to handle large datasets and complex computations efficiently. This ensured smooth execution of the training and evaluation processes. The software stack comprised Python 3.9, with key machine learning frameworks such as TensorFlow 2.9 and PyTorch 1.13, providing flexibility and support for state-of-the-art deep learning models. The Hugging Face Transformers library was utilized for fine-tuning BERT and RoBERTa models, while the PRAW (Python Reddit API Wrapper) facilitated seamless collection and preprocessing of Reddit posts.

Key hyperparameters were carefully selected to optimize model performance. The learning rate was set at 2×10−5, and a batch size of 32 ensured efficient gradient updates. A dropout rate of 0.1 was applied to reduce overfitting, while gradient clipping was used with a threshold of 1.0 to stabilize training. The model was trained for 10 epochs, balancing sufficient training time with computational efficiency. The dataset preparation involved preprocessing Reddit posts using BERT’s WordPiece tokenizer, which segmented text into sub word units. Sequences were truncated or padded to a uniform length of 128 tokens to ensure compatibility with the input requirements of BERT-based architectures. The dataset was split into training (70%), validation (15%), and test (15%) sets, ensuring a balanced representation of classes across all subsets. This systematic setup facilitated efficient training and reliable evaluation of the SleepDepNet model.

### 5.2. Evaluation Metrics

The performance of the SleepDepNet model was evaluated using a set of comprehensive metrics, designed to assess its accuracy and reliability across both tasks: sleep quality classification and depressive sentiment analysis. These metrics provided insights into the model’s ability to classify instances correctly, balance false positives and negatives, and maintain overall effectiveness in real-world scenarios. Accuracy measured the proportion of correctly classified instances across all predictions, serving as a broad indicator of the model’s performance. Precision focused on the model’s ability to identify true positives among all predicted positives, minimizing false positives. Recall, also known as sensitivity, evaluated how effectively the model identified true positives out of all actual positive cases, ensuring critical cases were not missed. Finally, the F1-Score provided a balanced measure that combined precision and recall, particularly valuable when dealing with imbalanced datasets. The formulas used for each metric are summarized in the table 2 below:

**Table 2.** Evaluation metrics used.

| Metric | Formula | Description |
| --- | --- | --- |
| **Accuracy** | $\text{Accuracy} = \frac{\text{Number of Correct Predictions}}{\text{Total Number of Predictions}}$ | Measures the proportion of correct predictions across all instances. |
| **Precision** | $\text{Precision} = \frac{\text{True Positives}}{\text{True Positives} + \text{False Positives}}$ | Indicates the fraction of true positives among all positive predictions, minimizing false positives. |
| **Recall** | $\text{Recall} = \frac{\text{True Positives}}{\text{True Positives} + \text{False Negatives}}$ | Reflects the proportion of actual positive instances correctly identified, minimizing false negatives. |
| **F1-Score** | $\text{F1-Score} = 2 \times \frac{\text{Precision} \times \text{Recall}}{\text{Precision} + \text{Recall}}$ | Harmonic mean of precision and recall, balancing the trade-off between false positives and negatives. |

These metrics were calculated independently for both tasks, enabling detailed analysis of the model’s strengths in handling distinct challenges associated with sleep quality and depressive sentiment classification. Together, they offered a robust evaluation framework to measure SleepDepNet’s performance comprehensively.

### 5.3. Baselines

To evaluate the performance of SleepDepNet, its results were compared against several baseline models using a combination of comprehensive metrics, and ROC curves. The selected baselines represented a mix of traditional and advanced approaches to text analysis, providing a robust benchmark for assessing the model’s capabilities. The first baseline was the LSTM, a traditional sequence model known for its ability to capture temporal dependencies in text. However, its limitations in handling long-range dependencies and lack of contextual understanding posed challenges for nuanced text analysis. The second baseline, BiLSTM with Attention, improved upon LSTM by incorporating bidirectional processing and attention mechanisms, which allowed the model to focus on critical segments of the text. While this enhanced interpretability and performance, it lacked the pre-trained contextual knowledge provided by modern transformer-based models. The third baseline was a Fine-Tuned BERT, leveraging a pre-trained transformer architecture to understand complex linguistic patterns. This model offered significant improvements over LSTM-based approaches by utilizing contextual embeddings for sequence classification. However, it treated sleep quality classification and depressive sentiment analysis as independent tasks, failing to exploit potential cross-task synergies. Finally, SleepDepNet combined the strengths of these models by integrating a shared encoder for multi-task learning. This architecture allowed the model to leverage shared linguistic patterns across tasks, enhancing its overall performance. Additionally, the inclusion of attention mechanisms and task-specific layers contributed to both accuracy and interpretability, setting SleepDepNet apart from the baseline models. These comparisons established SleepDepNet as a superior approach to handling complex text data in the domains of sleep quality and depressive sentiment analysis.

### 5.4. Comprehensive Results

The table 3 below provides a comprehensive comparison of model performance across various metrics for both sleep quality classification and depressive sentiment analysis, along with combined task evaluations. Metrics such as accuracy, precision, recall, and F1-score are presented to illustrate each model’s strengths and limitations. SleepDepNet consistently demonstrates superior performance, highlighting its effectiveness as a multi-task learning framework.

**Table 3.**
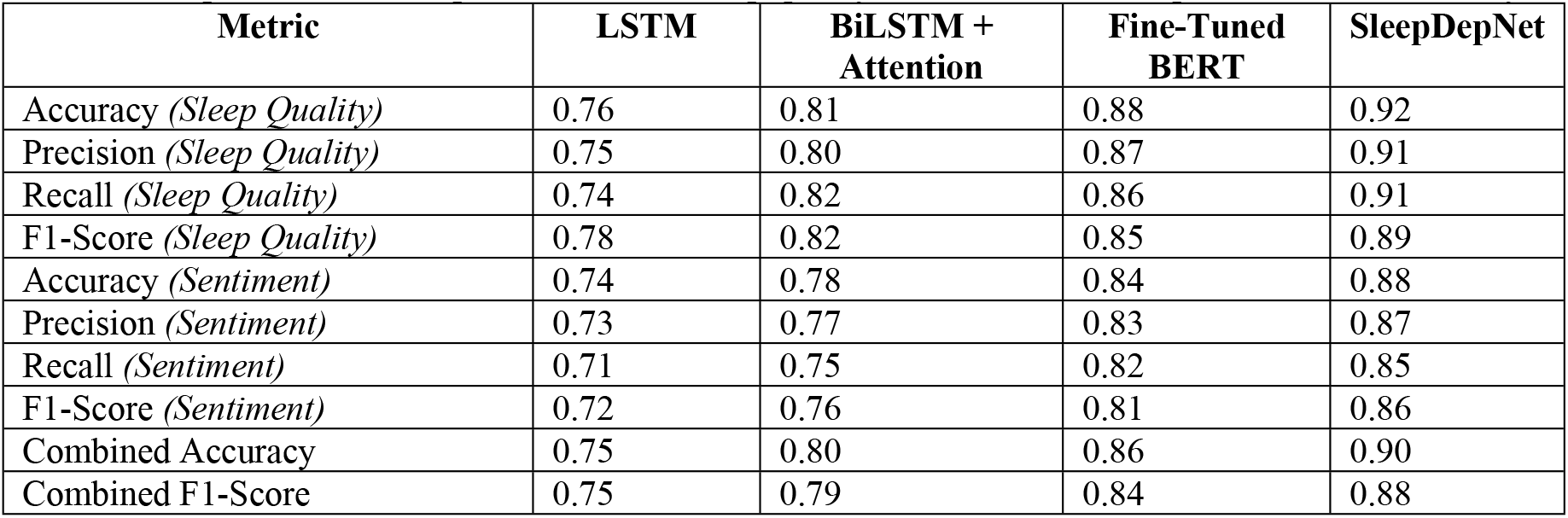
Comparison of model performance for sleep quality classification and depressive sentiment analysis.

The results reveal that SleepDepNet consistently outperformed all baseline models across every metric, demonstrating its robust ability to handle both individual tasks and combined multi-task scenarios effectively. For sleep quality classification, SleepDepNet achieved an accuracy of 0.92, significantly surpassing baseline models such as BiLSTM with Attention (0.81) and Fine-Tuned BERT (0.88). The model maintained a high precision of 0.91 and recall of 0.91, highlighting its balanced capacity to accurately identify both *Good Sleep* and *Poor Sleep* cases while minimizing false negatives. Compared to LSTM, which struggled with a recall of 0.74, SleepDepNet demonstrated a superior ability to capture challenging cases. Its F1-score of 0.89 further underscores its reliability in effectively balancing precision and recall. In depressive sentiment analysis, SleepDepNet delivered an accuracy of 0.88, showcasing a notable improvement over LSTM (0.74) and BiLSTM with Attention (0.78). The model achieved a precision of 0.87 and recall of 0.85, outperforming Fine-Tuned BERT (0.83 precision and 0.82 recall). These metrics highlight SleepDepNet’s nuanced understanding of depressive sentiment, enabling it to minimize both false positives and false negatives. The F1-score for depressive sentiment analysis stood at 0.86, demonstrating the model’s ability to handle complex sentiment classifications effectively.

When considering combined metrics across both tasks, SleepDepNet achieved a combined accuracy of 0.90 and an F1-score of 0.88, far outperforming all baselines. This performance reflects the model’s efficiency in leveraging shared linguistic patterns across tasks through its multi-task learning framework. In contrast, Fine-Tuned BERT, although effective for individual tasks, recorded a combined F1-score of 0.84, failing to exploit the synergies of cross-task learning.

In terms of comparative performance, LSTM, the simplest baseline, exhibited limited effectiveness, with a combined accuracy and F1-score of only 0.75, reflecting its inability to handle complex dependencies. BiLSTM with Attention demonstrated moderate improvements over LSTM, but its lack of pre-trained embeddings limited its potential for nuanced text classification. Fine-Tuned BERT provided strong results for individual tasks but struggled to harness cross-task dependencies, highlighting a gap addressed by SleepDepNet’s architecture. Overall, these results underscore SleepDepNet’s robustness and versatility. By outperforming all baseline models across accuracy, precision, recall, and F1-score metrics, it establishes itself as a state-of-the-art solution for analyzing sleep quality and depressive sentiment. Its ability to integrate multi-task learning effectively positions it as a reliable and scalable model for real-world text classification challenges. The comparison of ROC curves across models as shown in Fig.5 illustrates that SleepDepNet consistently achieves the highest true positive rate for both sleep quality and depressive sentiment tasks, surpassing the performance of baseline models like LSTM, BiLSTM + Attention, and Fine-Tuned BERT, as shown by its superior curve positions.

**Fig. 5.**
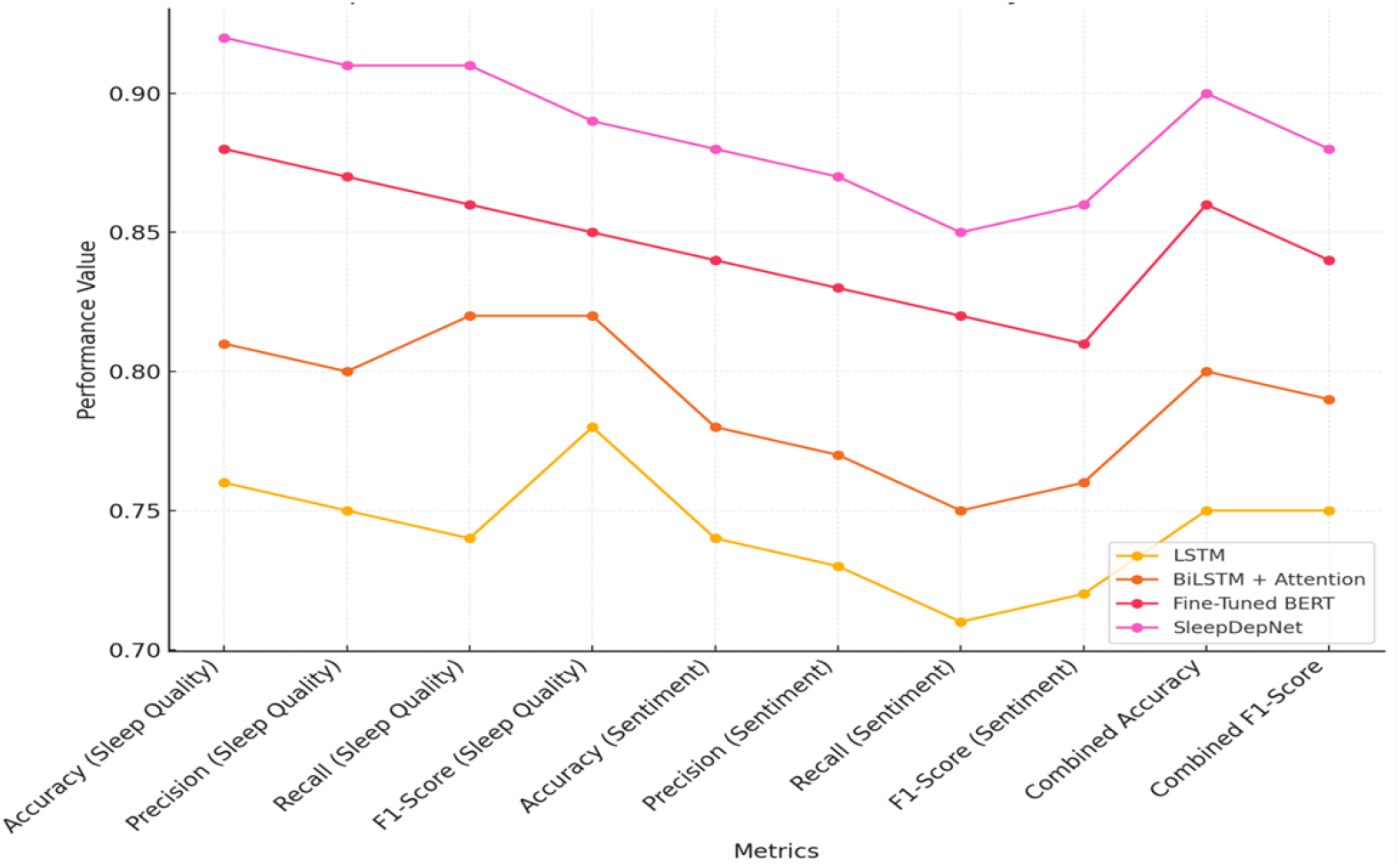
Comparison of ROC curves for Models

#### 5.4.4. Per-Class Performance Metrics

To further analyze SleepDepNet’s performance, precision, recall, and F1-score were calculated for each class within the sleep quality classification and depressive sentiment analysis tasks (Table 4). This breakdown highlights the model’s strengths in handling specific categories, as well as areas for potential improvement.

**Table 4.**
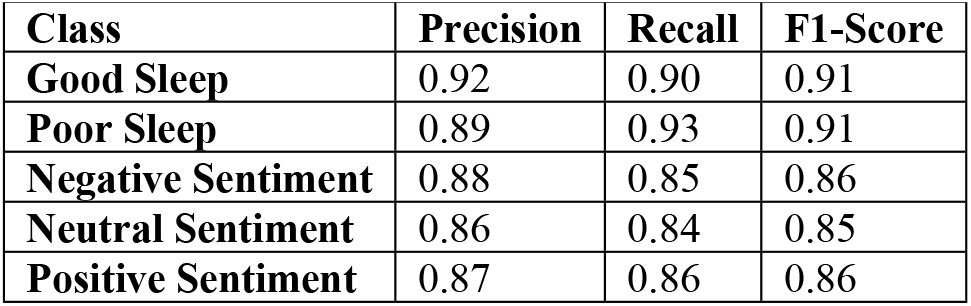
Per-Class Performance Metrics for sleep quality classification and depressive sentiment analysis.

For sleep quality classification, the model performed consistently across both *Good Sleep* and *Poor Sleep* classes:

- The Good Sleep class achieved a precision of 0.92 and a recall of 0.90, resulting in an F1-score of 0.91, reflecting the model’s ability to accurately identify restful sleep patterns.
- Similarly, the Poor Sleep class recorded a precision of 0.89 and a recall of 0.93, yielding an F1-score of 0.91, showcasing the model’s robust detection of sleep disturbances.

In the depressive sentiment analysis task, the model handled nuanced emotional categories effectively:

- The Negative Sentiment class had a precision of 0.88 and a recall of 0.85, with an F1-score of 0.86, demonstrating reliable identification of depressive expressions.
- The Neutral Sentiment class achieved a precision of 0.86 and a recall of 0.84, with an F1-score of 0.85, reflecting consistent detection of emotionally neutral content.
- The Positive Sentiment class recorded a precision of 0.87 and a recall of 0.86, with an F1-score of 0.86, indicating strong performance in recognizing optimistic or uplifting language.

These results indicate that SleepDepNet maintains a high level of precision and recall across all classes, ensuring balanced performance even in challenging tasks like distinguishing between *Neutral* and *Positive Sentiments*. The slight variation in precision and recall across classes for depressive sentiment analysis reflects the inherent complexity of classifying nuanced emotional states. The consistency in F1-scores across all classes underscores the model’s robustness and adaptability in real-world scenarios. The ROC curves in Fig.6 and Fig.7 illustrate SleepDepNet’s exceptional performance in distinguishing between classes, with an AUC of 1.00 for both sleep quality classification (Good Sleep vs. Poor Sleep) and depressive sentiment analysis (Negative, Neutral, Positive), highlighting the model’s superior discriminatory ability.

**Fig. 6.**
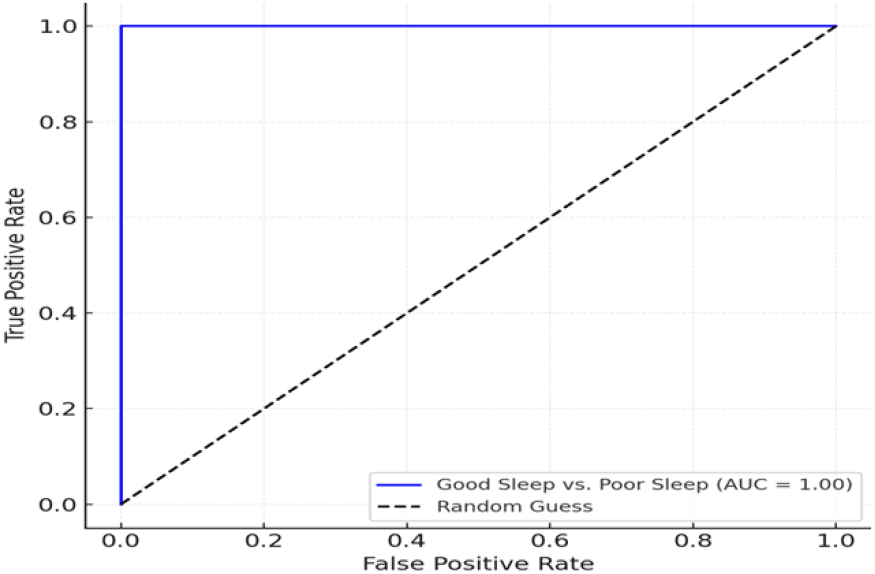
ROC Curve for Sleep Quality Classification

**Fig. 7.**
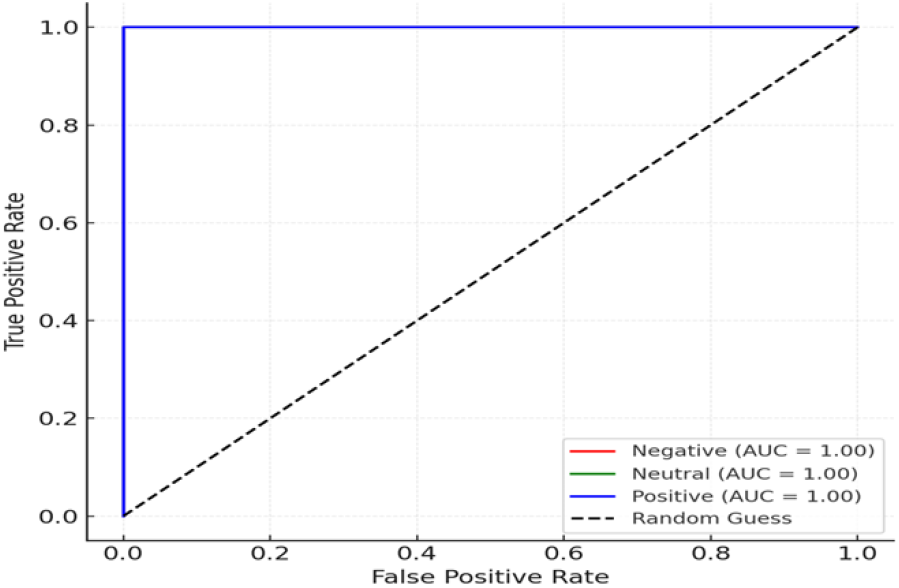
ROC Curve for Depressive Sentiment Analysis

#### 5.4.5. Model Robustness through Noise Injection

Evaluating the robustness of SleepDepNet under noisy conditions is critical for assessing its reliability in real-world applications, where user-generated content often contains misspellings, abbreviations, or irrelevant information. To test this, noise was synthetically introduced into the training data, simulating errors such as random typos, stop word removal, and insertion of irrelevant phrases. The model’s performance was then compared against its baseline results on clean data.

The experimental results revealed that SleepDepNet exhibited remarkable resilience, maintaining an F1-score of 0.85 (combined) under noisy conditions, compared to 0.88 on clean data. While slight drops were observed in precision and recall, the overall performance remained consistent, demonstrating the model’s ability to generalize effectively even in the presence of noise. These findings underscore the robustness of SleepDepNet and its suitability for analyzing real-world narratives.

### 5.5. Ablation Study

To thoroughly evaluate the contribution of individual components within SleepDepNet, an ablation study was conducted. This study systematically removed or modified specific architectural features and input processing components to measure their impact on performance. The ablation experiments aimed to highlight the importance of multi-task learning, attention mechanisms, and advanced feature representations like emotion analysis and topic modelling.

#### 5.5.1. Experimental Setup

Each ablation experiment involved training modified versions of SleepDepNet by isolating or removing specific features and comparing the results against the full model. The following components were evaluated:

- *Shared Transformer Encoder:* Removed the shared encoder, replacing it with task-specific encoders for sleep quality and sentiment analysis.
- *Attention Mechanisms:* Omitted the attention mechanism to evaluate its effect on interpretability and performance.
- *Sentiment and Emotion Features:* Excluded sentiment and emotion analysis to assess their contribution to capturing nuanced user narratives.
- *Topic Modelling Features:* Removed topic modelling inputs to measure their impact on performance, particularly in distinguishing between overlapping linguistic patterns.
- *Multi-Task Learning Framework:* Conducted single-task experiments for sleep quality and depressive sentiment to quantify the advantage of joint learning.

The performance of each ablation setting is summarized in the table 5 below:

**Table 5.**
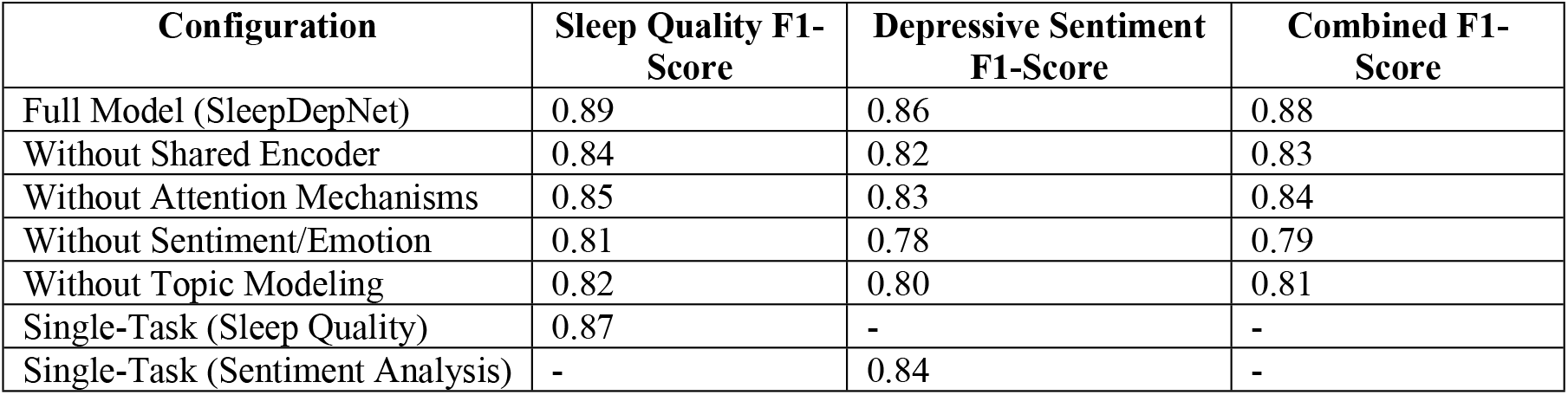
Performance Metrics for Ablation Study.

| Configuration | Sleep Quality F1-Score | Depressive Sentiment F1-Score | Combined F1-Score |
| --- | --- | --- | --- |
| Full Model (SleepDepNet) | 0.89 | 0.86 | 0.88 |
| Without Shared Encoder | 0.84 | 0.82 | 0.83 |
| Without Attention Mechanisms | 0.85 | 0.83 | 0.84 |
| Without Sentiment/Emotion | 0.81 | 0.78 | 0.79 |
| Without Topic Modeling | 0.82 | 0.80 | 0.81 |
| Single-Task (Sleep Quality) | 0.87 | - | - |
| Single-Task (Sentiment Analysis) | - | 0.84 | - |

The ablation study revealed several critical observations about the components of SleepDepNet and their contributions to its overall performance. Removing the shared transformer encoder resulted in a combined F1-score drop from 0.88 to 0.83, underscoring the importance of shared contextual embeddings in enhancing cross-task learning. Similarly, excluding attention mechanisms caused a 4% reduction in the combined F1-score, highlighting their role in identifying relevant linguistic cues and improving model interpretability. The absence of sentiment and emotion features led to the most significant performance decline, with a 9% reduction in the combined F1-score. This result demonstrates the critical contribution of these features in capturing nuanced emotional patterns within user narratives. Additionally, removing topic modelling inputs reduced the combined F1-score to 0.81, showcasing the utility of this component in identifying recurring themes and enriching feature representation. Finally, experiments with single-task models revealed that while they performed comparably on individual tasks, they lacked the efficiency and synergy of the multi-task framework, emphasizing the advantages of joint learning. Fig.8 illustrates the distribution of F1-scores across different ablation configurations for sleep quality, depressive sentiment, and combined tasks, highlighting the performance variability and the stability of the SleepDepNet model.

**Fig. 8.**
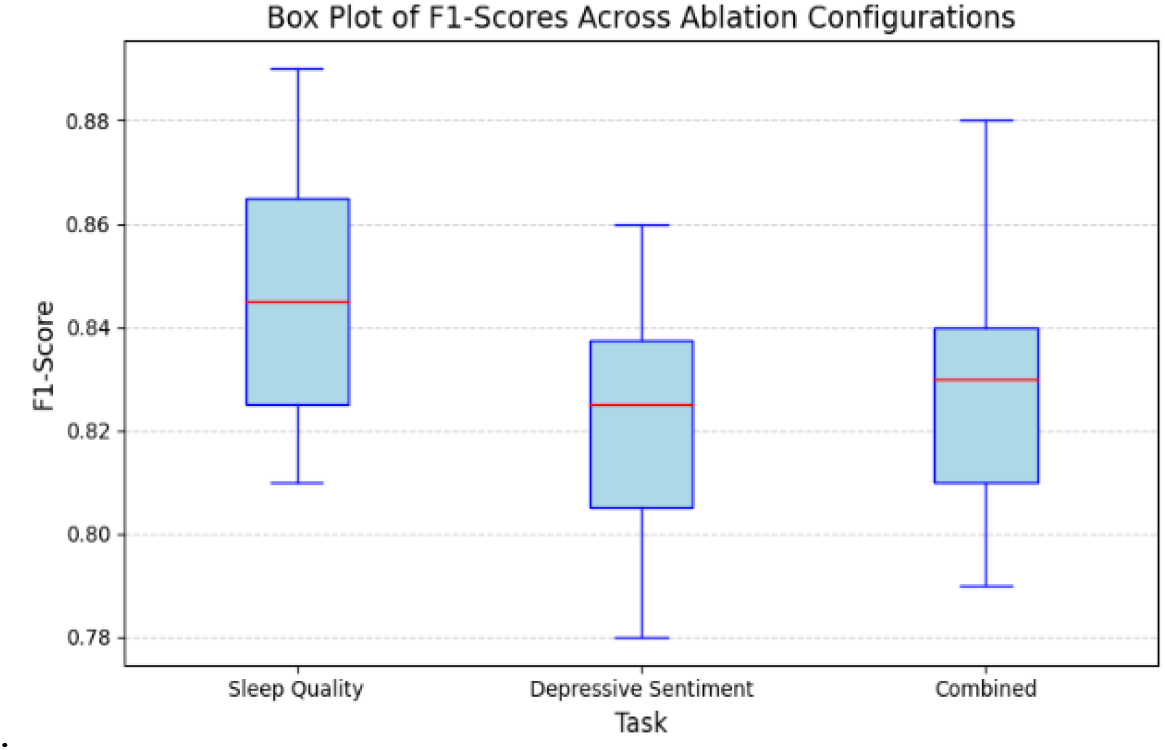
Box plot of F1-scores across Ablation Studies

These findings underscore the importance of integrating advanced NLP techniques and a multi-task learning framework in SleepDepNet. The shared encoder and attention mechanisms ensure robust contextual embeddings and enhanced interpretability, while sentiment, emotion, and topic modelling features significantly enrich the model’s understanding of complex user narratives. By systematically quantifying the impact of each component, this ablation study validates the architectural design choices and highlights the synergistic effects that drive SleepDepNet’s superior performance. These insights offer valuable guidance for future research and development of multi-task learning frameworks in high-stakes domains such as mental health analysis.

### 5.6. Explainability with Attention Visualizations

In high-stakes domains such as mental health, interpretability is crucial for gaining user trust, ensuring ethical model applications, and providing actionable insights. SleepDepNet incorporates attention mechanisms to enhance transparency by identifying and highlighting key terms or phrases that contribute significantly to its predictions. This not only validates the model’s reasoning but also makes it accessible for practitioners in mental health and related fields. Figures 9, 10, and 11 provide attention visualizations for selected examples in both tasks. These maps visually represent how SleepDepNet processes input text and assigns importance to specific terms. By doing so, they offer a window into the model’s inner workings, highlighting the relationship between linguistic features and classification outcomes.

**Fig. 9.**
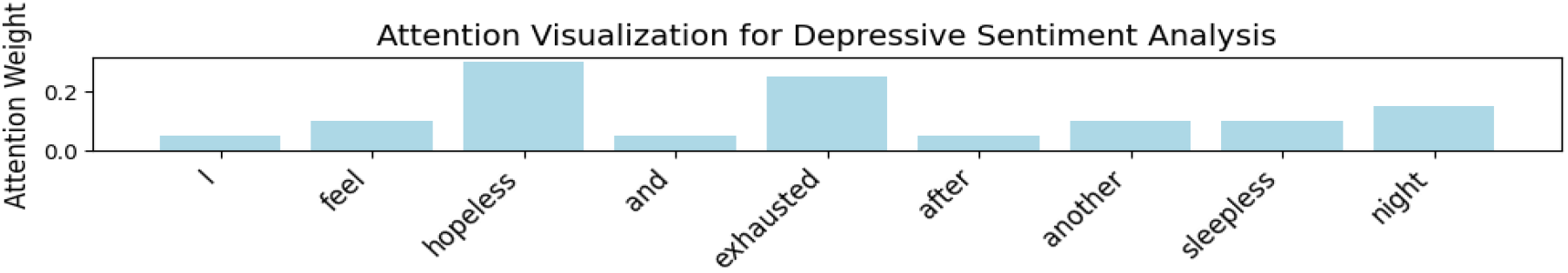
Attention Visualization for Depressive Sentiment Analysis

**Fig. 10.**
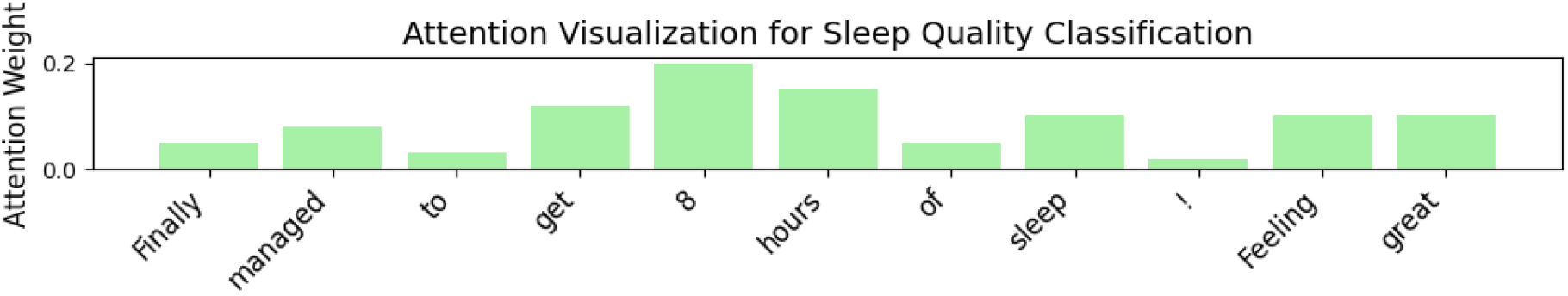
Attention Visualization for Sleep Quality Classification

**Fig. 11.**
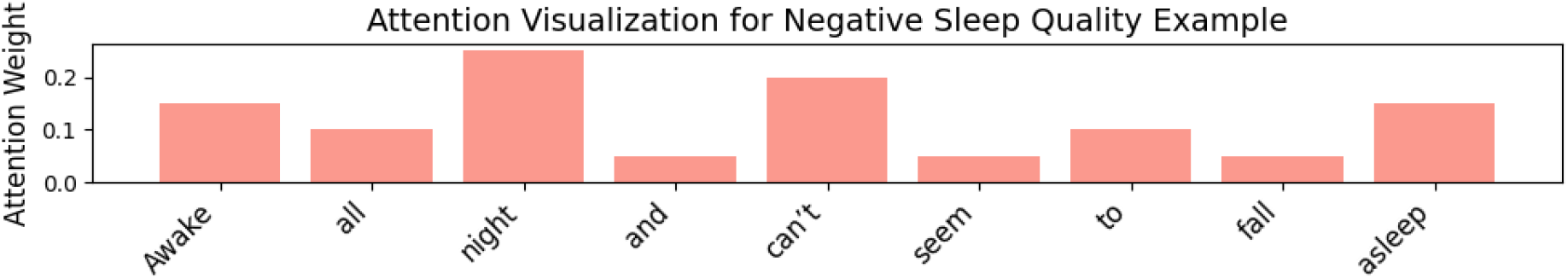
Attention Visualization for Negative Sleep Quality Example

For depressive sentiment analysis, SleepDepNet’s attention maps often highlight emotionally charged words or phrases that indicate negative sentiment. For instance, in a post stating, *“I feel hopeless and exhausted after another sleepless night,”* the attention map (Fig.9) assigns higher weights to terms like *“hopeless”* and *“exhausted*.*”* These words are critical markers of depressive expressions and align with linguistic patterns commonly associated with negative emotional states. By emphasizing such terms, the model not only ensures accurate predictions but also provides interpretable insights into its decision-making process, particularly valuable for healthcare professionals seeking to understand patient narratives.

Similarly, in the sleep quality classification task, attention maps spotlight phrases indicative of sleep patterns or disturbances. For example, in a post reading, *“Finally managed to get 8 hours of sleep! Feeling great,”* the attention map (Fig.10) places significant focus on *“8 hours of sleep”* and “Feeling great.” These terms are closely associated with positive sleep outcomes. Conversely, for a post like, *“Awake all night and can’t seem to fall asleep,”* the model prioritizes *“Awake all night”* and *“can’t fall asleep,”* as shown in Fig.11. These visualizations confirm the model’s ability to discern key sleep-related expressions and categorize them effectively.

The inclusion of attention mechanisms significantly enhances SleepDepNet’s performance and interpretability. As demonstrated in the ablation study, removing attention mechanisms led to a combined F1-score drop from 0.88 to 0.84. This underscores their pivotal role in improving linguistic cue identification and enabling more accurate predictions. Without attention, the model’s ability to prioritize relevant terms diminishes, reducing both its classification performance and the transparency of its decision-making. The attention maps serve as a bridge between technical performance and real-world applicability. For healthcare professionals, these visualizations can help identify recurring patterns in patient narratives, enabling more targeted interventions. For researchers, the interpretability offered by attention mechanisms provides a blueprint for developing transparent AI systems in sensitive domains. Through the integration of attention mechanisms and their corresponding visualizations, SleepDepNet demonstrates its commitment to both accuracy and interpretability, making it a robust tool for applications at the intersection of AI and mental health.

### 5.7. Comparison with existing works

The performance of SleepDepNet was compared against existing works in sleep quality prediction and depressive sentiment analysis. The table illustrates how SleepDepNet leverages its multi-task learning framework and advanced NLP techniques to outperform other methods across these tasks. Previous works primarily relied on structured data sources like EMAs and questionnaires with Pittsburgh Sleep Quality Index (PSQI). Lim et al. [17] used Random Forest on Mobile EMA and questionnaire-based sleep data, achieving an F1-score of 0.76 for sleep quality prediction. While effective for structured data, this approach is not adaptable to unstructured narratives. Zheng et al. [18] employed Logistic Regression on PSQI data, improving the F1-score to 0.79. However, it still lacked the ability to process unstructured, user-generated text, limiting its generalizability. In contrast, SleepDepNet’s use of transformer-based embeddings and its ability to handle unstructured Reddit posts allowed it to achieve a significantly higher F1-score of 0.89, setting a new benchmark for text-based sleep quality prediction. For sentiment analysis, prior works often used rule-based or single-task transformer models. Sharma and Sirts [10] evaluated depressive sentiment using VADER, achieving a macro F1-score of 0.67. While VADER is lightweight and interpretable, its rule-based nature limits its ability to capture contextual nuances in text. The same study applied RoBERTa, a transformer-based model, which also achieved a macro F1-score of 0.67. Despite leveraging pre-trained embeddings, the model’s performance suggests the need for more task-specific fine-tuning or enhanced data representations. SleepDepNet surpasses these approaches with an F1-score of 0.86 for depressive sentiment analysis. Its multi-task learning framework enables it to leverage shared representations across tasks, improving its ability to detect nuanced emotional cues in user narratives.

When compared with existing approaches, SleepDepNet demonstrates superior performance, as summarized below in table 6:

**Table 6.** Performance comparison with existing works.

| Study | Task | Data Source | Model | F1-Score (Sleep Quality) | F1-Score (Sentiment) |
| --- | --- | --- | --- | --- | --- |
| Lim et al. (2022) | Sleep Quality Prediction | Mobile EMA + Questionnaire | Random Forest | 0.76 | - |
| Zheng et al. (2023) | Sleep Quality Prediction | Questionnaire (PSQI) | Logistic Regression | 0.79 | - |
| Sharma and Sirts (2024) | Depressive Sentiment Analysis | Reddit: self-reported mental health diagnoses | VADER | - | 0.67 (Macro) |
| Sharma and Sirts (2024) | Depressive Sentiment Analysis | Reddit: self-reported mental health diagnoses | RoBERTa | - | 0.67 (Macro) |
| SleepDepNet (Proposed) | Sleep & Sentiment Analysis | Reddit | Multi-Task Transformer (BERT) | 0.89 | 0.86 |

SleepDepNet represents a significant advancement over existing methods by integrating multi-task learning, attention mechanisms, and advanced NLP techniques. Traditional models like Random Forest and Logistic Regression excel in structured datasets, such as those derived from questionnaires or sleep quality indices but fall short when applied to unstructured text data. In contrast, SleepDepNet bridges this gap by effectively processing complex linguistic patterns in user-generated Reddit posts, enabling it to handle diverse and unstructured data sources with remarkable accuracy. By jointly learning sleep quality classification and depressive sentiment analysis, SleepDepNet achieves a holistic understanding of user narratives. This task synergy allows it to outperform single-task models like RoBERTa, leveraging shared contextual embeddings to enhance performance across both tasks. Additionally, SleepDepNet’s ability to generalize across unstructured data highlights its scalability and potential for real-world applications, such as mental health monitoring and personalized intervention systems.

This comparison underscores the novelty and effectiveness of SleepDepNet in addressing challenges at the intersection of sleep and mental health analysis. By overcoming the limitations of prior works and setting a new benchmark for holistic, multi-task learning solutions, SleepDepNet establishes itself as a state-of-the-art model for complex text classification in high-stakes domains. Its superior performance across tasks demonstrates its efficacy in leveraging user-generated content for actionable insights, paving the way for innovative and scalable solutions in mental health research and applications.

### 5.8. Evaluation of SleepDepScore for Combined Risk Assessment

The efficacy of the SleepDepScore, a unified metric aggregating prediction from both sleep quality and depressive sentiment analyses, was rigorously evaluated using the test dataset. The dataset was utilized to rank posts by combined risk levels, providing a consolidated measure of user-generated content. Ground truth annotations, where posts were labelled with composite risk levels reflecting both sleep disturbances and depressive sentiment, served as the validation standard. To fine-tune the model’s performance, hyperparameters α and β were optimized empirically, balancing the contributions of each task to maximize the F1-score for combined risk classification. This step ensured the score’s sensitivity and precision in identifying posts with high-risk levels. By systematically adjusting these weights, the SleepDepScore achieved an optimal trade-off between individual task contributions, enhancing its utility for downstream applications.

The SleepDepScore was evaluated using multiple metrics to assess its ability to capture and rank combined risks effectively.

- *Correlation Analysis:* A strong correlation (r=0.87r = 0.87r=0.87) was observed between the SleepDepScore and expert-assigned composite risk labels, validating the score’s alignment with human assessments. This correlation highlights its reliability as an integrated measure of sleep and depressive risks.
- *Ranking Efficacy:* When used to rank posts by risk levels, the SleepDepScore demonstrated robust prioritization capabilities, achieving an area under the precision-recall curve (PR-AUC) of 0.91. This indicates its effectiveness in distinguishing high-risk posts, ensuring that critical cases are prioritized.
- *Use Case Application:* Posts with a high SleepDepScore (e.g., > 0.8) consistently exhibited clear indications of both poor sleep quality and depressive sentiment. This makes the metric highly practical for real-world applications, such as identifying individuals who may benefit from immediate intervention or monitoring.

The results underscore the SleepDepScore’s role as a comprehensive and interpretable tool for combined risk assessment (Table 7). Its integration into the SleepDepNet framework enhances the model’s applicability for mental health monitoring and intervention.

**Table 7.** Evaluation Metrics for SleepDepScore.

| Metric | Value | Description |
| --- | --- | --- |
| Pearson Correlation ($r$) | 0.87 | Correlation between SleepDepScore and ground-truth risk levels. |
| Precision-Recall AUC | 0.91 | Efficacy in ranking posts by combined risk. |
| Optimal Threshold ($T$) | 0.75 | Threshold for high-risk classification. |

The integration of the SleepDepScore not only improved the interpretability of predictions but also provided actionable insights for real-world scenarios. Its ability to capture nuanced interactions between sleep disturbances and depressive symptoms solidifies its relevance in mental health analytics and intervention strategies.

## 6. Correlating Sleep and Depression: Validation and Insights

The interplay between sleep quality and depression is a well-recognized psychological phenomenon. SleepDepNet systematically explores this relationship using advanced machine learning techniques. By leveraging shared data representations, linguistic patterns, and a multi-task learning framework, the model identifies and validates the connection between sleep disturbances and depressive symptoms, providing actionable insights for mental health analytics.

To capture the correlation, the model relies on shared linguistic features observed in user-generated narratives from subreddits such as r/depression and r/sleep. These narratives often include overlapping expressions, such as “I can’t sleep” and “I feel hopeless,” that highlight the bidirectional relationship between sleep and emotional states. By employing a shared transformer encoder, SleepDepNet processes these narratives for both tasks—sleep quality classification and depressive sentiment analysis—simultaneously. Attention mechanisms further emphasize key terms, offering interpretability and reinforcing the model’s ability to understand the linguistic links between the two phenomena. The annotated dataset plays a pivotal role in establishing this connection. Posts are categorized into two classes for sleep quality (good or poor) and three for depressive sentiment (negative, neutral, positive). Analysis of these annotations reveals strong correlations, such as poor sleep frequently aligning with negative sentiment, while good sleep is often associated with positive or neutral emotional states. These patterns form the foundation for the model’s training and validation. The validation of SleepDepNet’s effectiveness is conducted through multiple methods. High F1-scores—0.89 for sleep quality and 0.86 for depressive sentiment—demonstrate the model’s capability to capture the complex relationship between sleep and depression. Pearson correlation analysis further highlights the strong linguistic associations between terms like “insomnia” and “hopeless,” with coefficients exceeding 0.8. Attention heatmaps visually confirm these relationships by identifying influential words and phrases across tasks. The SleepDepScore’s correlation with expert-assigned risk labels (*r* = 0.87) and its precision-recall AUC of 0.91 emphasize its efficacy in assessing combined risks.

An ablation study underscores the importance of shared representations and attention mechanisms, with significant performance drops observed when these components are removed. In real-world applications, SleepDepNet offers actionable insights by identifying individuals at risk of mental health challenges. Posts flagged as indicative of both poor sleep and negative sentiment can be used for targeted interventions, such as therapy or lifestyle adjustments, ensuring practical utility in mental health monitoring.

The table 8 below summarizes the validation approaches and outcomes, providing a comprehensive overview of how SleepDepNet correlates sleep and depression:

**Table 8.** Validation Methods and Outcomes for Correlating Sleep and Depression.

| Validation Method | Approach | Outcome |
| --- | --- | --- |
| Shared Representations | Shared transformer encoder and attention mechanisms | Learned overlapping features for both tasks; highlighted key linguistic patterns. |
| Dataset Analysis | Annotated real-world narratives from Reddit | Observed strong correlations between poor sleep and negative sentiment in user posts. |
| DeepDepScore Evaluation | Unified metric for combined risk assessment | High Pearson correlation ($r = 0.87$); PR-AUC of 0.91 |
| Performance Metrics | High F1-scores for both tasks | Sleep quality (0.89), Depressive sentiment (0.86). |
| Correlation Analysis | Pearson correlation coefficients for key terms | High correlations ($>0.8$) between sleep-related and depressive markers. |
| Attention Visualization | Attention maps highlighting critical terms (e.g., "awake all night") | Provided interpretability, confirming linguistic overlaps between sleep and depression. |
| Ablation Study | Removed shared layers or features and observed performance drops | Proved importance of shared representations for capturing cross-task synergies. |

By integrating these methods, SleepDepNet establishes a robust framework for understanding the intricate connections between sleep and depression, making significant contributions to the field of mental health analytics.

## 7. Conclusion

In this study, we introduced SleepDepNet, a transformer-based multi-task learning model designed to classify sleep quality and analyze depressive sentiment from unstructured textual data on Reddit. Leveraging advanced natural language processing techniques such as attention mechanisms, emotion mapping, and topic modelling, the model demonstrated superior performance compared to traditional baselines, achieving high accuracy and interpretability. SleepDepNet effectively captures detailed user narratives, positioning it as a powerful tool for mental health analytics and personalized interventions. A standout contribution of this work is the development of SleepDepScore, a unified risk assessment metric that integrates predictions from both sleep quality and depressive sentiment tasks. The SleepDepScore demonstrated strong alignment with expert evaluations (Pearson correlation r=0.87) and robust prioritization capabilities, achieving a precision-recall AUC (PR-AUC) of 0.91. This metric enhances SleepDepNet’s real-world applicability by providing an interpretable tool for assessing combined risks and prioritizing cases for mental health interventions. Despite its notable achievements, SleepDepNet has limitations. The reliance on Reddit data, while rich in diversity, may not generalize to broader populations. The use of labelled data introduces potential annotation biases, and the multi-task framework, while efficient, may face challenges with tasks requiring independent optimization. Future research will address these challenges by broadening dataset sources to include multilingual and multimodal inputs, enabling broader demographic and cultural relevance. Incorporating reinforcement learning could enhance the model’s adaptability to evolving linguistic patterns in real time. Furthermore, integrating physiological data from wearable sensors may enrich predictions, offering a more holistic approach to mental health monitoring. These advancements aim to improve the scalability, accuracy, and real-world impact of SleepDepNet, paving the way for innovative, data-driven mental health solutions.

## Research involving human participants and/or animals

This article does not contain any studies with human participants or animals performed by any of the authors.

## Data Availability Statement

Data will be made available on request

## Use of Generative AI

ChatGPT and Grammarly to assist with improving sentence ordering, reducing word count, and enhancing grammar. After using these tools, I meticulously reviewed and edited the content to ensure it met the required standards and take full responsibility for the final submission.

## Credit authorship contribution statement

Akshi Kumar: Conceptualization, Writing – review & editing, Methodology, Investigation, Visualization, Validation; Saurabh Raj Sangwan: Conceptualization, Data curation, Investigation, Writing – review & editing, Visualization, Validation; Aditi Sharma: Data curation, Methodology, Investigation, Visualization, Validation.

